# Multi-omics clustering analysis carries out the molecular specific subtypes of thyroid carcinoma: implicating for the precise treatment strategies

**DOI:** 10.1101/2024.02.25.24303184

**Authors:** Zhenglin Wang, Qijun Han, Xianyu Hu, Xu Wang, Rui Sun, Siwei Huang, Wei Chen

**Author notes:** These authors contributed equally to the study (Zhenglin Wang, Qijun Han, Xianyu Hu). **Correspondence to:** Prof. Wei Chen, Department of General Surgery, The First Affiliated Hospital of Anhui Medical University Hefei, Anhui Province 230022, China, Jixi Road 218, Hefei, Anhui Province 230022, P.R. China.

## Abstract

**Background:** Thyroid cancer is the most prevalent endocrine malignancy, Recent classifications highlight the importance of molecular characteristics in TC, including BRAF, TERT, and RET fusion gene mutations, which are crucial for diagnosis, prognosis, and targeted therapy. This study aims to explore molecular subtypes of TC to identify new biomarkers and improve patient selection for targeted therapies.

**Methods:** This study utilized multi-omics data from the TCGA-THCA dataset and additional cohorts (GSE29265, GSE33630, GSE54958, GSE65074) involving a total of 539 patients. Various data types, including DNA methylation, gene mutations, mRNA, LncRNA, and miRNA expression, were analyzed. The study employed consensus clustering algorithms to identify molecular subtypes and used various bioinformatics tools to analyze genetic alterations, signaling pathways, immune infiltration, and responses to chemotherapy and immunotherapy. The statistical significance was established at P < 0.05.

**Results:** Two prognostically relevant thyroid cancer subtypes, termed CS1 and CS2, were identified. CS2 was associated with a poorer prognosis of shorter progression-free survival times (P < 0.001). CS1 exhibited higher copy number alterations but lower tumor mutation burden (TMB) than CS2. Notably, CS2 showed higher TMB and cytolytic activity scores, suggesting a potential for higher immunogenicity. Different pathway activations were observed between subtypes, with CS2 showing activation in cell proliferation and immune-related pathways. Drug sensitivity analysis indicated CS2’s higher sensitivity to cisplatin, doxorubicin, paclitaxel, and sunitinib, whereas CS1 was more sensitive to bicalutamide and FH535. The different activated pathways and sensitive to drugs for subtypes were further validated in external cohort. After dimensionality reduction, five genes of CXCL17, LCN2, MUC1, SERPINA1, and SLC34A2 were validated that can distinguish subtypes across pan-cohorts. 24 paired tumor and adjacent normal tissues by immunohistochemical staining further show the prognostic value of CXCL17 for advanced thyroid cancer.

**Conclusion:** The study revealed two distinct molecular subtypes of thyroid cancer with significant implications for prognosis, genetic alterations, pathway activation, and treatment response. These findings underscore the potential of multi-omics approaches in enhancing personalized medicine in thyroid cancer.

## Introduction

Thyroid cancer is the most common malignant tumor of endocrine system^1^, represents approximately 95% of all endocrine tumors, accounting for 3–4% of all cancers, with an incidence rate ranking ninth among all cancers in 2020^2^. In China, the incidence rate of TC, reaching 14.56/100,000 has ranked the seventh in overall malignant tumors^3^. Specifically, the incidence of thyroid cancer is rising rapidly in the female population, compared with the national cancer epidemiological statistics in 2012, the latest data show that the incidence of thyroid cancer in women has risen from seventh to third place^3,4^.The World Health Organization histological classification distinguishes four main types of DTC derived from epithelial thyroid cells in which is the most common pathological type, accounting for about 80%-90% of all thyroid cancers ^5,6^. With appropriate treatment, patients with DTC carry an overall excellent prognosis. Surgical treatment is the main treatment for thyroid cancer, followed by endocrine therapy, radionuclide therapy, radiotherapy and targeted therapy in some cases^7^. Surgical treatment includes traditional neck open surgery, endoscope-assisted neck traceless surgery, robot-assisted surgery and so on^8^. Surgical treatment is still the main treatment for rare follicular carcinoma and medullary carcinoma^9^. Undifferentiated thyroid carcinoma is highly malignant, and a few patients have the opportunity of surgery. Some patients may have certain effect after radiotherapy and chemotherapy, but overall the prognosis is poor and the survival time is short^10^.

In the WHO 5th edition of the Classification of Thyroid Tumors, malignant tumors of follicular cell origin of the thyroid gland are classified according to molecular characteristics and aggressiveness. Papillary thyroid carcinoma (PTC) with multiple histological subtypes is a BRAF-like malignant tumor. Infiltrating encapsulated follicular subtype PTC and thyroid follicular carcinoma are RAS-like malignancies^11^. By single-cell RNA sequencing, a distinct BRAF-like B subtype has been uncovered that is predominantly dedifferentiated-like thyrocytes, enriched cancer-associated fibroblasts, worse prognosis and promising prospect of immunotherapy ^12^. The discovery of BRAF, TERT and RET fusion gene mutations provides a basis for the auxiliary diagnosis, tumor biological behavior, prognosis judgment and efficient and precise treatment of thyroid cancer patients. Several drugs have been developed that inhibit signaling kinases or oncogenic kinases (BRAFV600E, RET/PTC), such as those related to VEGFR/PDGFR^13,14^. Tyrosine kinase receptors are involved in cell proliferation and angiogenesis of thyroid cancer. Tyrosine kinase inhibitors (TKIs) are emerging as new treatments for DTC, MTC, and anaplastic TC (ATC), and can induce clinical response and stable disease ^15–17^.

Based on the molecular characteristics of thyroid cancer, more studies are required to identify potentially effective targeted therapies. Therefore, we collected and analyzed multi-omics data on thyroid cancer. Through our research, we will discover new biomarkers and more precisely select patients which may benefit from specific targeted therapy.

## Methods

### Data collection

Multi-omics data from TCGA-THCA dataset were downloaded for the molecular subtyping for TC patients, which included the omics data of DNA methylation, gene mutations, and extensive RNA sequence (mRNA, LncRNA, miRNA). The expression data of transcriptomics and clinical features were obtained via the “TCGAbiolinks” package. Gene symbols annotation was performed along with our prior study^18^. miRNA expression profile and DNA methylation 450 matrix were downloaded from UCSC Xena (https://xenabrowser.net/datapages), and somatic mutations from the cBioPortal (https://www.cbioportal.org/). After merging the all available data in different patients, 390 patients from the TCGA-THCA cohort remained for the molecular-specific subtypes. Another 149 TC patients were also collected, from GSE29265, GSE33630, GSE54958, and GSE65074 cohorts^19–22^. Batch effects are systematic technical bias unrelated to the biological state introduced when samples are processed and measured in different batches.. To make the GEO-sourced cohorts more similar in nonbiological parameters, the ComBat algorithms was employed to remove the batch effects between the four cohorts based on the R package “sva”. The data of the four GEO cohorts were merged into a new cohort as the validated cohorts, while the TCGA-THCA was used as the training cohort. (**Fig. S1**), while the TCGA-THCA cohort was used as the training cohort in the subsequent analysis. The detailed information of enrolled cohorts was listed in **Table 1**.

**Table1.**
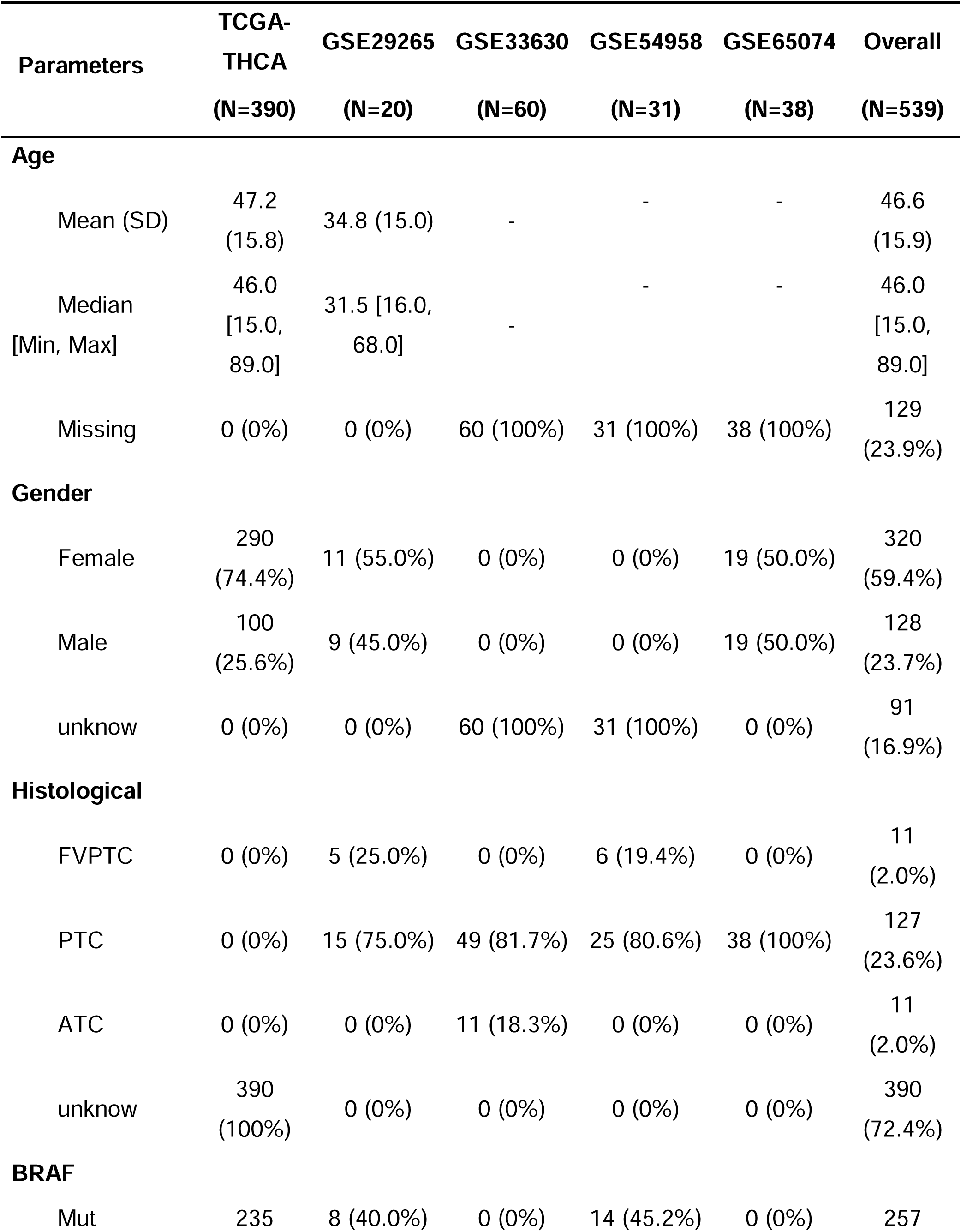
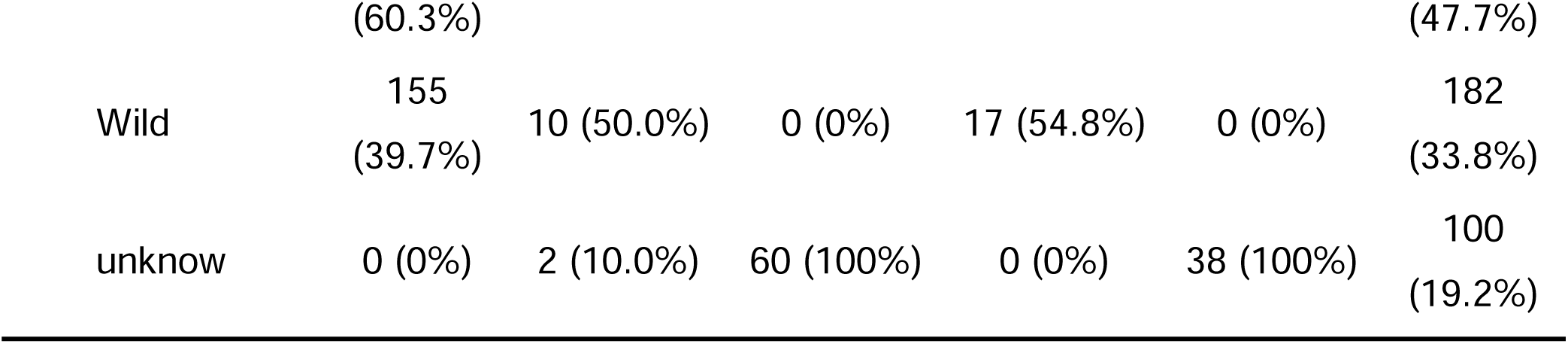
The clinicopathological features of enrolled cohorts.

#### Construction of multi-omics molecular classification

The core algorithms for consensus clustering were from recently published R package “*MOVICS*”^23^, and the molecular subtypes were confirmed using the enrolled multi-omics data. Univariate Cox regression was firstly performed to filtrate the progression-free survival (PFS) associated factors, which included miRNA, LncRNA, mRNA, and DNA methylation CpG sites, by the threshold *P* value < 0.05. Mutant genes were selected with a mutant frequency higher than 10%. Algorithms of clustering prediction index (CPI) ^24^ and Gaps-statistics ^25^ were conducted to find the appropriate number of subtypes with the multi-omics data. Ten clustering algorithms which included iClusterBayes, moCluster, CIMLR, IntNMF, ConsensusClustering, COCA, NEMO, PINSPlus, SNF, and LRA were obtained from the “*MOVICS*”. We preset the optimal clusters for the ten clustering algorithms in to separate the patients into different clusters, and further combined the results by consensus ensembles to identify the robust subtypes. Eventually, we calculated the average silhouette width for each subtype to test the cluster quality.

### Analysis of genetic alteration features between different subtypes

The tumor mutation burden (TMB), defined by the number of mutations per million bases, was calculated, as well as Fraction genome altered (FGA) which are referred to as the percentage of the genome affected by copy number gains (FGG) or losses (FGL). We obtained the total neo antigens and cytolytic activity (CYT) scores from a previous study ^26^. The neoantigens were predicted by analyzing tumor-specific mutational frameshifts, splices, gene fusion, endogenous retroelement, and other criteria. The copy number segment data were downloaded from FireBrowse (http://firebrowse.org/) and visualized by the “maftools” package in R.

### Evaluation of the activated of signaling pathways and immune infiltration

Fifty HALLMARK terms of each patient were annotated using the single-sample gene set enrichment analysis (ssGSEA) using the “GSVA” package. The enrichment score (ES) was the primary result of the gene set enrichment analysis, reflecting the degree to which a gene set is overrepresented in the top or bottom of a ranked list of genes. A positive ES indicates that the gene set is enriched at the top of the ranking list, while a negative ES indicates that the gene set is enriched at the bottom of the ranking list. The normalized enrichment score (NES) is the primary statistic for examining gene set enrichment results. The false discovery rate (FDR) is the estimated probability that a gene set with a given NES represents a false-positive finding. The significance threshold of the enrichment score for a single gene set was defined by *P*-value < 0.05. From a previous study, we obtained the gene sets for immune and stromal signatures ^27^ and calculated the NES score to compare the difference in immune-activated status among the different subtypes. The mean enrichment score for each subtype was used for visualization by a heatmap. For the altered activated transcriptional regulatory networks (regulons), we performed the “RTN” analysis ^28^, and several chromatin remodeling-associated genes were also analyzed to reveal the potential regulons^29^. Details of regulon calculation pointed out in a prior study^30^. The BRAF-like and RAS-like subtypes were generated in a prior study^31^, and we used them to compare with the subtyping in the current study.

### Evaluation of responses to chemotherapy and immunotherapy

To evaluate the individual likelihood of responsiveness to immunotherapy, we obtained a melanoma cohort contained patients who received anti-CTLA-4 or anti-PD-1 checkpoint inhibition therapy or not, as well as specific gene sets with 795 genes were obtained ^32^. We performed Subclass mapping analysis to compare the similarity between risk groups with the immunotherapy subgroups and predict the responders to anti-CTLA-4 or anti-PD-1 therapy ^33^. The response to chemotherapy for each patient was predicted based on the Genomics of Drug Sensitivity in Cancer (GDSC) database. Accordingly, five commonly used drugs, cisplatin, doxorubicin, paclitaxel, sunitinib, bicalutamide, and FH535 were selected. The responses to the above-mentioned five drugs were compared by estimating the half-maximal inhibitory concentration (IC_50_) of the samples by ridge regression.

### Statistical analysis

All analyses were performed using the R software v4.1.2 (http://www.r-project.org). Continuous data were compared using the student’s t-test or Wilcoxon test, and the Chi-square test was used to compare the distribution of categorical variables. The connection between the two factors was assessed by calculating Pearson correlation coefficient. To reproduce the classification in the external validation cohort, nearest template prediction (NTP) analysis was performed via the top 200 specific marker genes in the TCGA-THCA cohort ^34^. The survival difference of PFS between groups was described by K-M curves. The log-rank test was used to estimate the survival analysis, and HR and 95% CI were calculated using the Cox model. The independent prognostic effect of the multi-omics classification was calculated using multivariate cox regression analysis. *P <* 0.05 was considered a statistically significant difference.

## Results

### Identification of two prognosis-relevant subtypes

Based on the results in the CPI analysis and Gaps-statistics analysis, we found the scores were all at the highest values when the number of clusters is tw o (**Fig. S2**), therefore, we pre-set two clusters to perform the ten-omics integrative clustering and then combined via ensemble consensus. Two robust subtypes were finally identified and termed as CS1 and CS2 (**Fig. 1B**). Based on the multi-omics data from the TCGA-THCA cohort, we visualized the multi-omics features in the two subtypes and annotated the top ten items of each omics in the right (**Fig. 1A**). The two subtypes exhibited distinct clinical outcome in survival analysis, and CS2 had a significantly shorter PFS time, indicating a poor-prognosis phenotype (*P* < 0.001, **Fig. 1C**). To further access the cluster reliability, silhouette analysis was conducted, and the average silhouette width of 0.62 and 0.63 manifested the appropriate cluster configuration (**Fig. 1D**). We compared the distribution of clinical and pathological features in two groups. More White patients were belonged to CS2 group (*P* < 0.001), and with the advanced tumor stage of stage III and IV group (*P* < 0.001), as well as the number of livings with tumor (*P* = 0.001, **Table 2**). Interestingly, the distribution of patients with alive or dead clinical outcome in two subtypes showed no difference (*P* = 0.043), alghough they shared the diverse RFS status. Moreover, we evaluated the independent prognostic value of multi-omics subtype, after adjusting age, tumor stage, gender and tumor status, the multi-omics subtype still presented independent RFS prognostic value (HR: 3.763, 95% CI: 1.269-11.16, *P* =0.017, **Table 3**).

**Figure 1.**
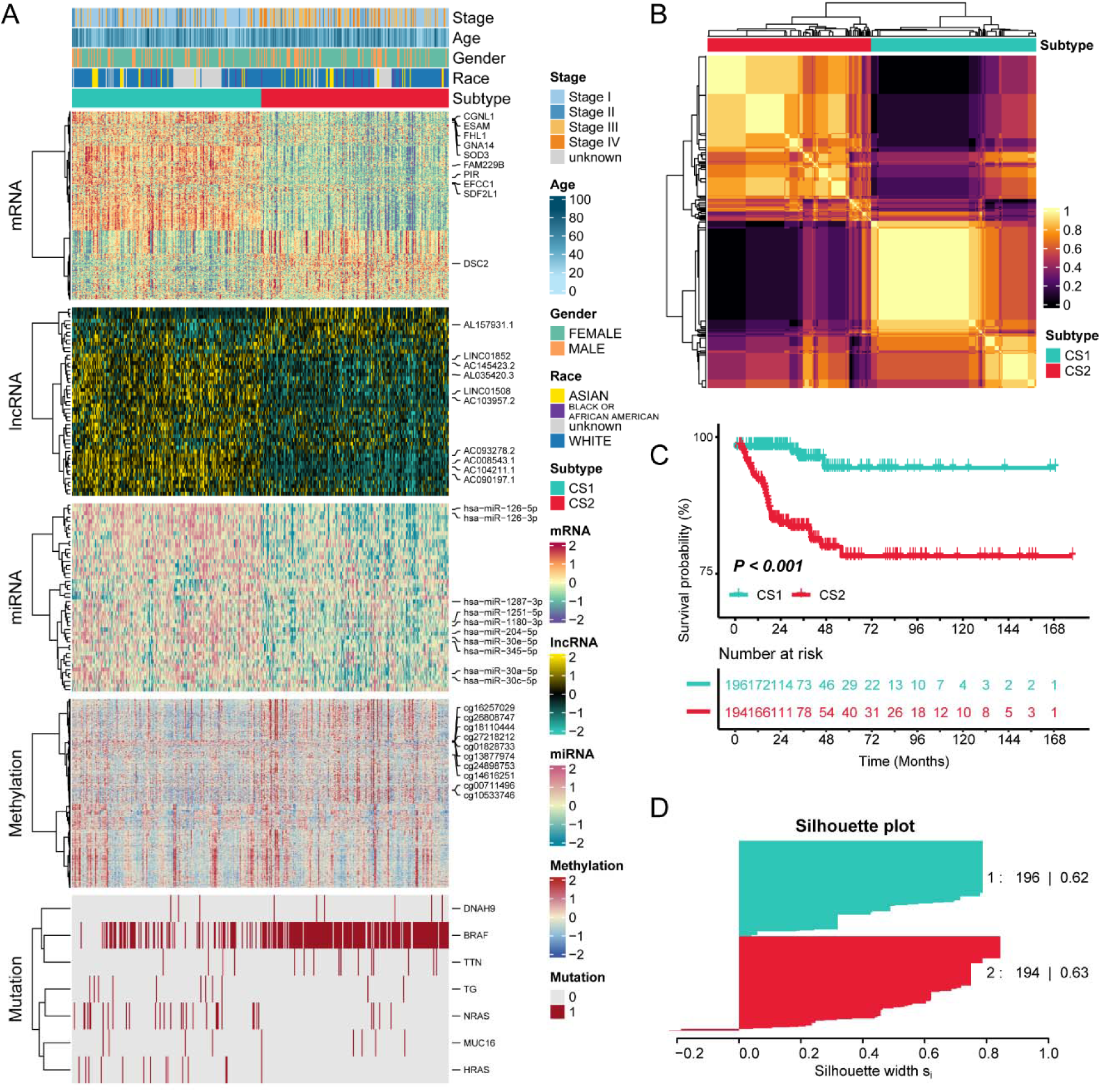
Recognition of the muti-omics subtypes based on multi-omics data. (A) The different omics features of mRNA, lncRNA, miRNA, methylation, and mutation between CS1 and CS2 subtypes, and the top ten items in each omics were listed on the right. (B) Consensus clustering for muti-omics subtypes based on the ten algorithms; (C) K-M curves for the two subtypes; (D) Silhouette analysis for the two subtypes, and the silhouette width close to 1 indicated the appropriate cluster configuration.

**Table 2.** Diverse distribution of clinical features in CS1 and CS2 of TCGA-THCA.

|  | CS1 | CS2 | P value |
| --- | --- | --- | --- |
| **Age, years** | 46.28 ± 15.56 | 48.16 ± 15.96 | 0.237 |
| **Gender (%)** |  |  |  |
| Female | 149 (76.0%) | 141 (72.7%) | 0.523 |
| Male | 47 (24.0%) | 53 (27.3%) |  |
| **Race (%)** |  |  |  |
| Asian | 16 (8.2%) | 21 (10.8%) | <0.001* |
| Black or African American | 7 (3.6%) | 11 (5.7%) |  |
| Unknown | 56 (28.6%) | 19 (9.8%) |  |
| White | 117 (59.7%) | 143 (73.7%) |  |
| **Stage (%)** |  |  |  |
| Stage I | 126 (64.3%) | 98 (50.5%) | <0.001* |
| Stage II | 30 (15.3%) | 12 (6.2%) |  |
| Stage III | 28 (14.3%) | 53 (27.3%) |  |
| Stage IV | 11 (5.6%) | 30 (15.5%) |  |
| Unknown | 1 (0.5%) | 1 (0.5%) |  |
| **Vital status (%)** |  |  |  |
| Alive | 191 (97.4%) | 185 (95.4%) | 0.403 |
| Dead | 5 (2.6%) | 9 (4.6%) |  |
| **Tumor status (%)** |  |  |  |
| Tumor free | 188 (96.9%) | 163 (86.7%) |  |
| With tumor | 6 (3.1%) | 24 (12.8%) |  |
| Unknown | 0 (0.0%) | 1 (0.5%) | 0.001* |

\*, $P < 0.05$ .

**Table 3.** Prognostic value of multi-omics subtype and clinicopathological features assessed by multivariate Cox regression analysis.

| **Features** | **HR** | **95%CI** | **P value** |
| --- | --- | --- | --- |
| **Age, years** | 0.988 | 0.951-1.030 | 0.549 |
| **Stage** |  |  |  |
| II vs. I | 0.763 | 0.151-3.850 | 0.743 |
| III vs. I | 1.952 | 0.546-6.79 | 0.309 |
| IV vs. I | 1.615 | 0.356-7.32 | 0.535 |
| **Gender** |  |  |  |
| Male vs. Female | 1.266 | 0.567-2.82 | 0.565 |
| **Tumor status** |  |  |  |
| With tumor vs. Tumor free | 15.84 | 7.228-34.71 | <0.001* |
| **Cluster** |  |  |  |
| CS2 vs. CS1 | 3.763 | 1.269-11.16 | 0.017* |

\*, P < 0.05.

### CS1 exhibited higher copy number alterations (CNAs) while CS2 exhibited higher TMB

Genetic alteration is an important factor in the depiction of the biological behavior of tumors. Therefore, we compared the differences of CNA, TMB, neo antigen, CYT and gene mutations among CS1 and CS2. We observed that the CS1 subtype presented higher FGA, FGL (all *P* < 0.05, **Fig. 2A**) but lower TMB (*P* = 7.5e – 03, **Fig. 2B**) than CS2 subtype. The higher TMB in CS2 might be related to high immunogenicity. The genetic instability of tumor cells contributes to the development of non-synonymous mutations, and the expressed products are unique antigens, which are now referred to as neoantigens^35^. Regrettably, there is no statistical difference in neoantigen was observed between CS1 and CS2 (*P* = 0.33, **Fig. 2C**). In addition, cytolytic activity (CYT) score was defined based on the expression level of granzyme A and perforin. It was established to quantify the immune cytolytic activity and has been identified to relate to the clinical outcomes of many tumors^36,37^. We found CS2 subtype had a higher CYT score than CS1 (*P* = 0.00014, **Fig. 2D**), indicating high CYT score correlated to a poor prognosis in thyroid cancer. We compared the mutated genes among multi-omics classification and listed the top four in **Fig. 2E**. CS1 exhibited higher mutation frequencies of NRAS, HRAS, and EIF1AX, and CS2 had more BRAF mutations (all *P* < 0.05).

**Figure 2.**
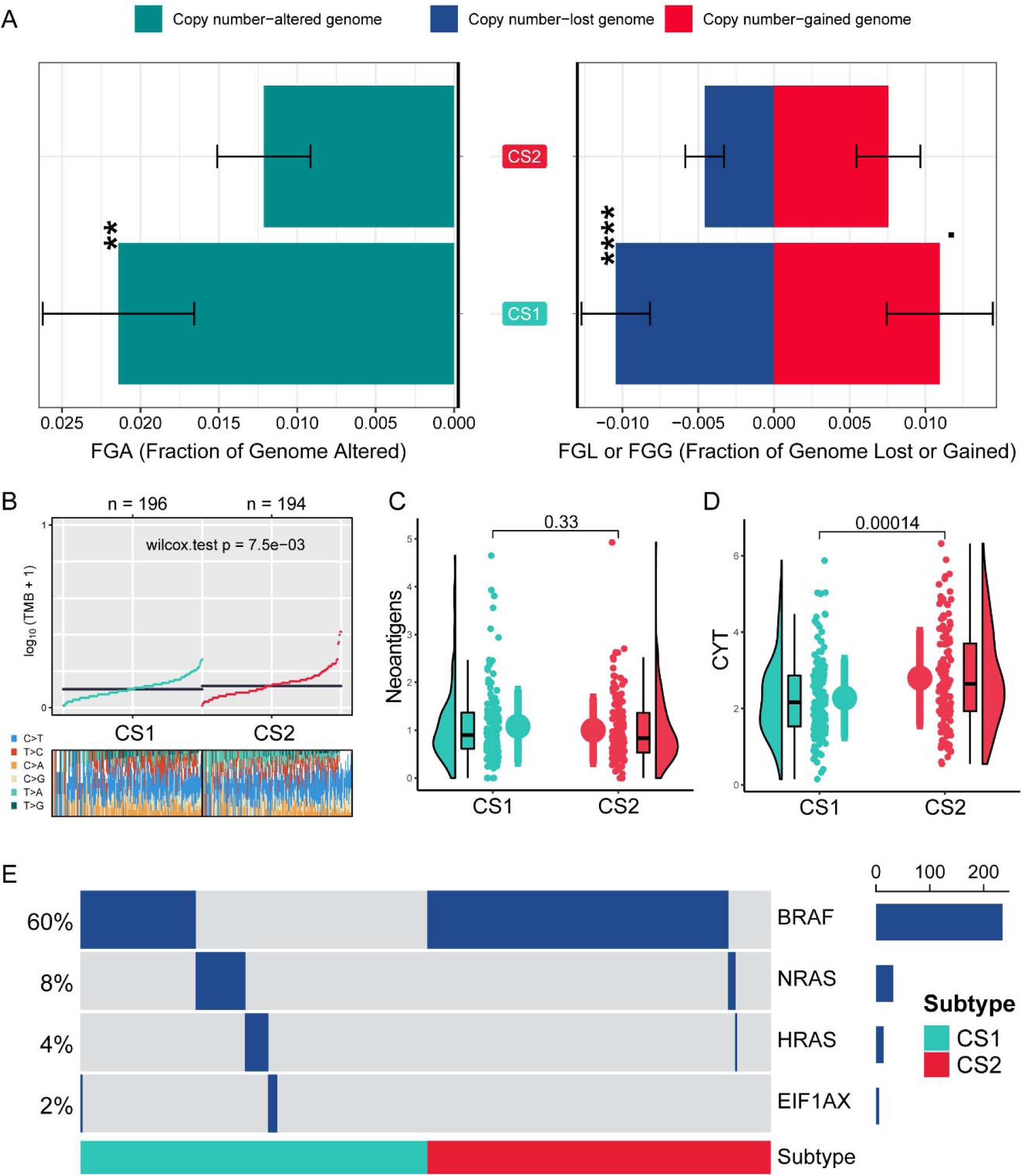
Distinct genetic alteration among muti-omics subtypes. (A) Comparison of copy number alterations between CS1 and CS2 subtypes, including the fraction of genome, altered, fraction of genome lost, and the fraction of genome gained. (B) Comparison of tumor mutation burden among muti-omics subtypes. (C) Comparison of total neoantigens, which were predicted by analyzing tumor-specific mutational frameshifts, splices, gene fusion, endogenous retroelement, and other criteria. (D) Different cytolytic activity between CS1 and CS2 subtypes and the score was defined based on the expression level of granzyme A and perforin level. (E) The top four characteristic gene alterations in CS1 and CS2 subtypes.

### Distinct pathway activation and chemotherapy response were observed among muti-omics subtypes

We explored the contrasting activated biological process, aim to detailly illustrate the specific molecular characteristics. CS2 subtype presented high activation of cell proliferation pathways, such as G2M checkpoint, MYC targets, E2F targets, as well as several traditional tumor-associated pathways, such as epithelial-mesenchymal transition, angiogenesis, KRAS signalling, P53 pathway, and hypoxia pathway. We also revealed the abnormally activated immune relevant pathways, such as IFN-α, IFN-γ, inflammatory response, IL6 JAK STAT3 signalling, IL2 STAT signalling, and TGF-β signalling (**Fig. 3A**). Upregulation of these proliferation pathways explained the poor prognosis of CS2 patients, and the high immune relevant pathways in CS2 were consistent with its high CYT score. As for CS1 subtype, we observed the activated pathways of oxidative phosphorylation, pancreas β cells, adipogenesis, fatty acid metabolism and WNT/β-catenin signalling (**Fig. 3A**). The enhancements of beta cells and adipogenesis signalling contributed to the decreased free fatty acids, thereby hindering the progression of CS1 tumor^38,39^. The characterized pathways of the CS1 and CS2 subtypes were further studied separately by tree plot via “clusterProfiler” package. The most relevant pathways of CS1 were organic acid transport, vascular transport, transport across the blood-brain, barrier, vascular process circulatory system, potassium ion transport, second-messenger-mediated signalling, steroid metabolic process, regulation of membrane potential, and theroid hormone relevant pathways (**Fig. 3B**). In CS2 subtype, the most notable pathway was immune-relevant, such as positive regulation of T cell activation, antigen processing, presentation of exogenous antigen and immunocytes migration (**Fig. 3C**), which was consistent with the results form HALLMARK 50 tumor pathways.

**Figure 3.**
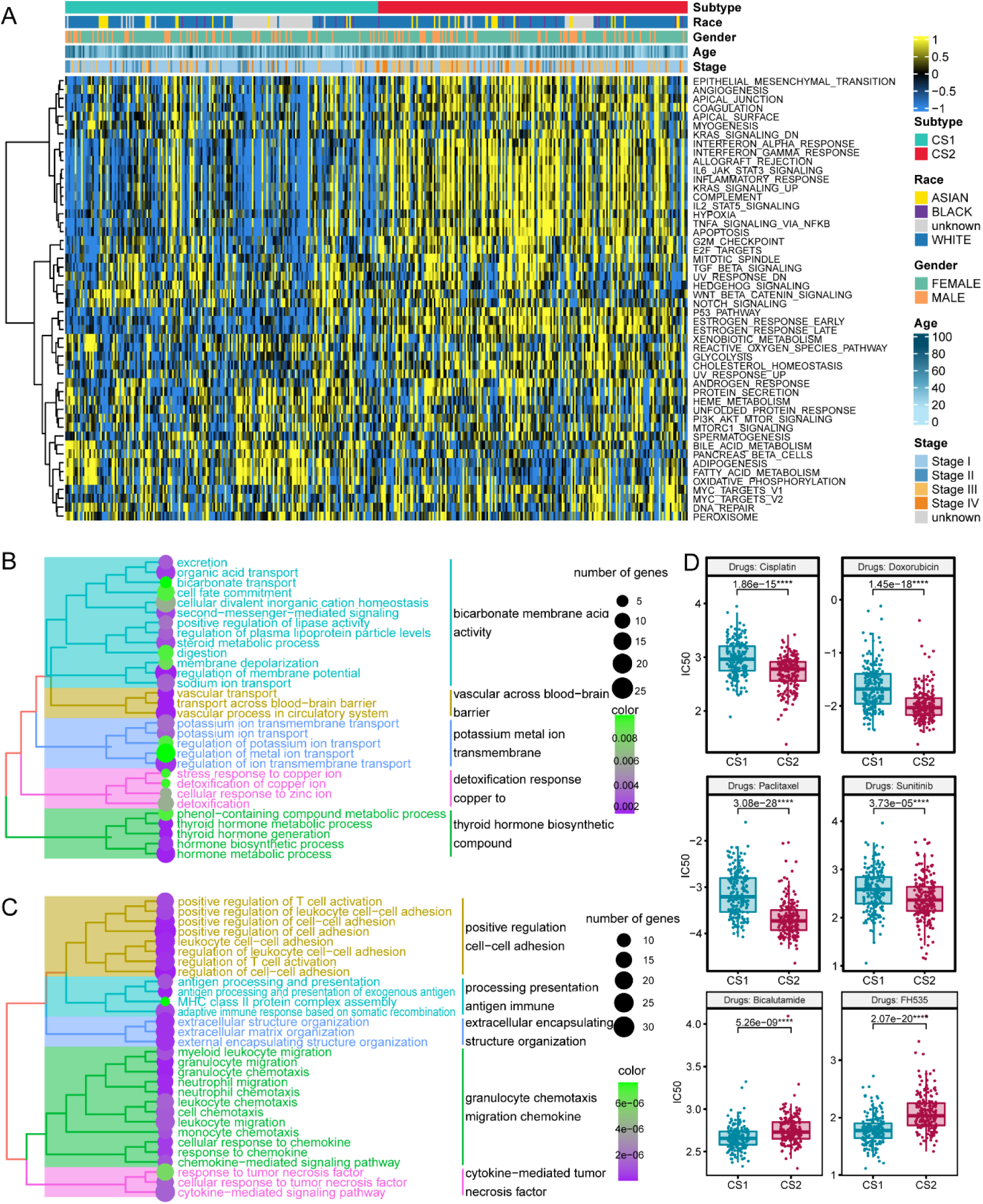
Different molecular features and prediction of chemotherapy response. (A) Different activated tumor-associated pathways among muti-omics subtypes; (B) Characteristic signaling pathways in CS1 subtype; (C) Characteristic signaling pathways in CS2 subtype. (D) Response prediction to different chemotherapeutic drugs, including cisplatin, doxorubicin, paclitaxel, sunitinib, bicalutamide, and FH535. The therapeutic efficacy was evaluated via IC50, and high IC50 represented a poor outcome.

In addition, the two subtypes were evaluated for treatment response to different chemotherapy agents. For cisplatin (*P* = 1.86e-51), doxorubicin (*P* = 1.45e-18), paclitaxel (*P* = 3.08e-28), and sunitinib (*P* = 3.73e-05), CS2 had lower IC50 compared with the CS1 subtype, indicating patients in CS2 had a higher sensitivity to these drugs. For bicalutamide (*P* = 5.2e-09) and FH535 (a small-molecule Inhibitor for Wnt/β-Catenin) (*P* = 2.07e-20), significantly lower IC50 were observed in the CS1 subtype, which means the CS1 patients would be more sensitive to these two drugs (**Fig. 3D**).

### Multi-omics classification can distinguish responders to immunotherapy and had a connection with BRAF-RAS classification

As mentioned above, there were substantial variations in immune status among the two subtypes, and CS2 exhibited higher CYT and activated immune-relevant pathways. Therefore, we focused our further analyses on the different immune features between CS1 and CS2 subtypes. We performed the immune cell infiltration landscape analysis and found that CS2 tumors contained more infiltration of immune cells, such as activated CD8 T cell, activated B cell, immature B cell, mast cell, activated dendritic cell, natural killer T cell, T follicular helper cell, effector memory B cell and activated CD4 T cell (**Fig. 4A**). Meanwhile, CS2 tumors presented higher expression level of PD-1, PD-L1, PD-L2, and CTLA-4 as compared with CS1 subtype (all *P* < 0.05, **Fig. 4B**). The predictive value of high TMB, activated CD8 T cell infiltration and PD-1 expression level in anti-PD-1 therapy has been reported in many studies, which suggested CS2 subtype might be a candidate for such treatment^40,41^. In SubMap analysis, we found CS2 patients were potential responders to anti-PD-1 or anti-PD-L1 therapy, but not CS1 patients, based on the data from two independent PD-1/L1 treated cohort (**Fig. 4C-D**). In addition, we elucidated activated transcriptional factors for CS1 and CS2 subtypes to explore their different molecular mechanisms, and the results were showed by the combined gene expression levels of their direct targets, which are also named transcriptional factor regulons^42^. As showed in **Fig. 4E**, CS1 had highly activated ESR2, PPARG, AR, RARB, FGFR1, RXRA, RXRB, and FGFR3 regulons, and CS2 presented highly activated ERBB3, GATA3, ESR1, RARG, FOXA1, PGR, TP63, FOXM1, and HIF1A regulons.

**Figure 4.**
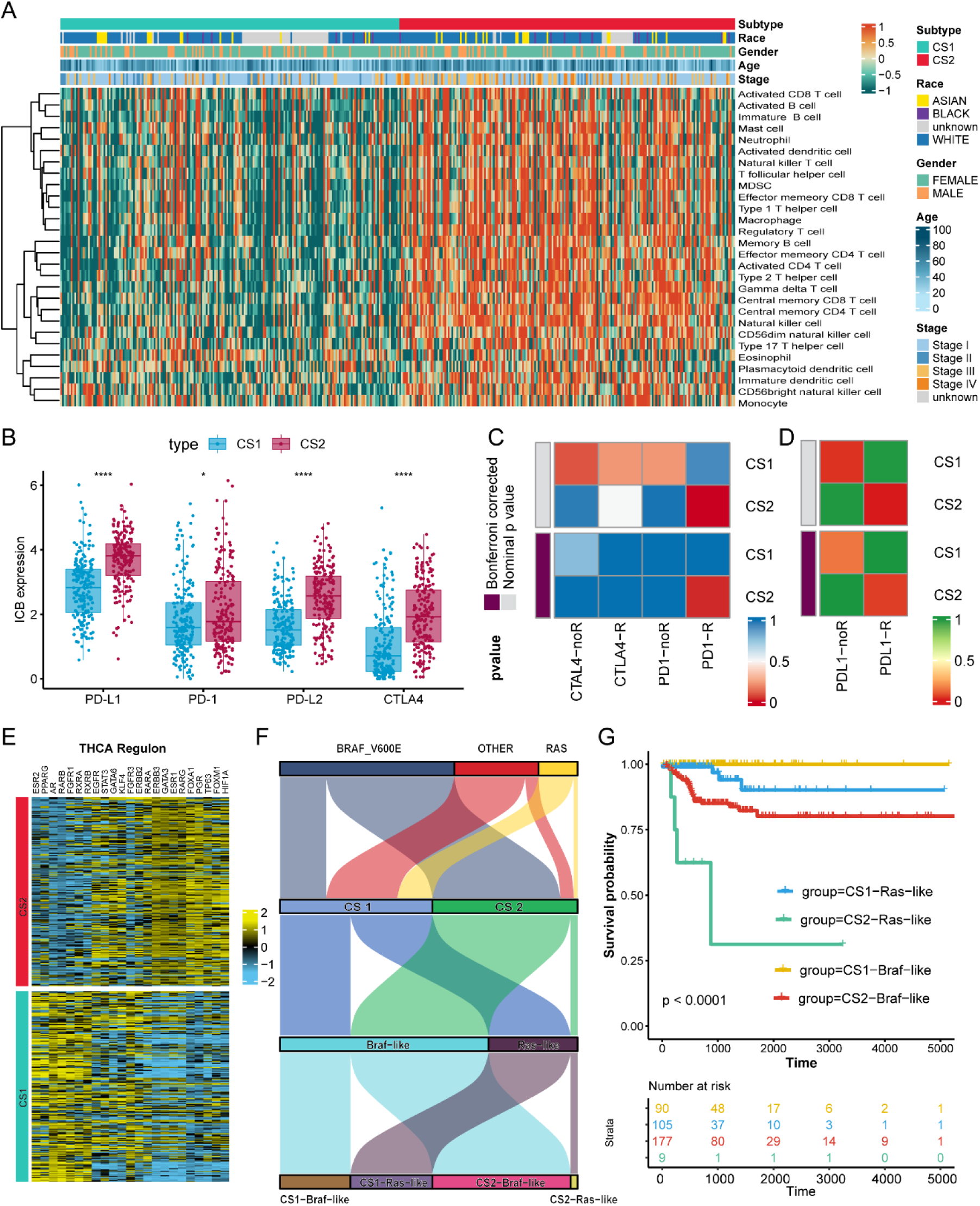
The different immunocyte infiltration, sensitivity to immunotherapy, and regulons among muti-omics subtypes, and the connections between muti-omics classification and BRAF^V600E^-RAS classification. (A) Distinct immunocyte infiltration landscape among multi-omics subtypes. (B) CS1 and CS2 subtypes showed variable expression levels of PD-L1, PD-1, PD-L2, and CTLA-4. (C) SubMap analysis showed the responders to anti-PD-1 or anti-CTLA-4 therapy, the dark red square represented the positive response. (D) SubMap analysis revealed the responders to anti-PD-L1 therapy. (E) The different regulons transcriptional regulatory networks (regulons) between CS1 and CS2 subtypes, the results were showed by corresponding targeted genes. (F) The Sankey diagram showed the connection degree between muti-omics subtypes, BRAF-like or RAS-like clusters, and their driven genes. (G) The different prognoses of the four redefined subtypes.

We compared the Multi-omics classification with the BRAF^V600E^-RAS classification and showed their connections via Sankey diagram^43^(**Fig. 4F**). For BRAF^V600E^ mutant tumors, most belonged to CS2 subtype, and CS1 subtype contained more RAS mutation. Therefore, vast majority of inferred RAS-like tumors belonged to the CS1 subtype, while most inferred Braf-like tumors in CS2 subtype. In addition, combined with CS1, CS2, BRAF-like, and RAS-like subtypes, we further refined thyroid cancer into CS1-BRAF like, CS1-RAS like, CS2-BRAF like, and CS2-RAS like subtypes. Among the four subtypes, CS2-RAS like tumour showed the worst prognosis, CS1-BRAF like tumour had the longest RFS, CS1-RAS like and CS2-BRAF like were with the moderate outcome (*P* < 0.0001, **Fig. 4G**). It indicated that a minority of patients had the worst prognosis, which is consistent with the actual situation^44^.

### Extra validation of the molecular features in GEO-combined cohort

To apply the muti-omics classification in GEO-combined cohort, we selected the top 500 genes with the highest expression in each subtype for grouping (**Fig. S3**). Patients in GEO-combined cohort were further divided into CS1 and CS2 by the 500 template genes. The similar subtype-specific pathways were also observed in the two subtypes. High activation of pancreas beta cells, adipogenesis, and fatty acid metabolism was also observed in CS1 subtype. In the CS2 subtype, we observed MYC targets V1 and V2, mitotic spindle, TNGA signalling via NF-kB, IL2 STAT5 signalling, KRAS signalling, and immune-relevant pathways, like interferon-αresponse, interferon-γ, and response complement pathways (**Fig. 5A**). Furthermore, similar regulons were found observed in the transcriptional regulatory network. CS1 contained activated AR, RARB and RXRA, and CS2 contained activated FOXM1, HIF1A, RARA, RARG, ERBB3, EGFR, and STAT3(**Fig. 5B**). We further investigated the characterized pathways of the CS1 and CS2 subtypes by “clusterProfiler”, respectively. As noted above, the CS1 contained bicarbonate transport and thyroid hormone biosynthetic and metabolic processes (**Fig. 5C**); the CS2 subtype contained various immune-related pathways, like lymphocyte-mediated immunity, adaptive immune response based on somatic recombination, antigen presentation processing, and immunocytes migration (**Fig. 5D**).

**Figure 5.**
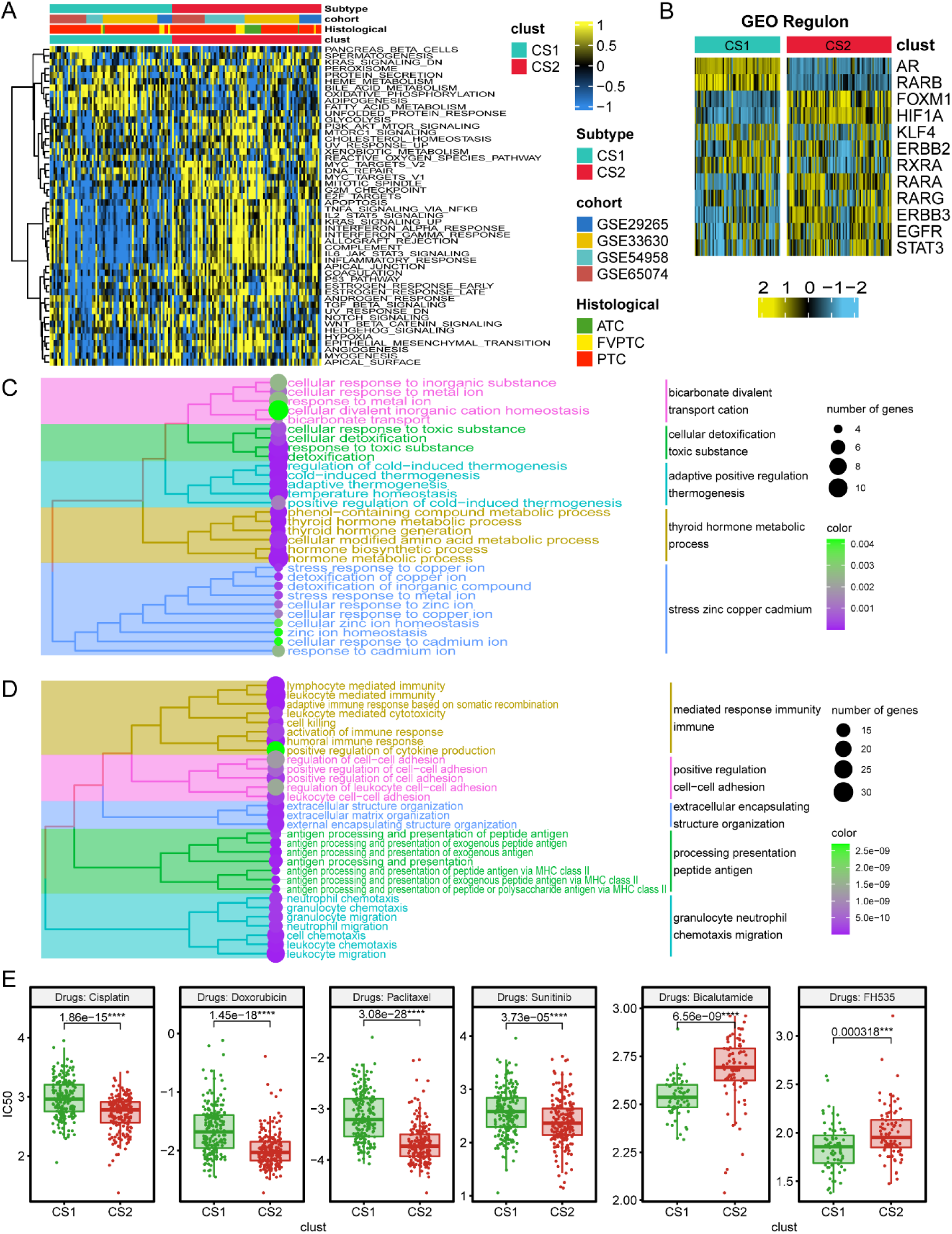
The different activated signalling pathways among muti-omics subtypes and response to chemotherapy in GEO-combined cohort. (A) CS1 and CS2 subtypes showed variable tumor-associated signalling pathways. (B) Regulon analysis in GEO-combined cohort. (C) Characteristic signaling pathways in CS1 subtype. (D) Characteristic signaling pathways in CS2 subtype. (E) Different response to cisplatin, doxorubicin, paclitaxel, sunitinib, bicalutamide, and FH535.

Based on the GEO-combined cohort, we tested the therapeutic value of the chemo drugs. CS1 subtype exhibited higher sensitive to bicalutamide (*P* = 6.56e - 09) and FH 535 (*P* = 0.000318), while CS2 showed favourable response to cisplatin (*P* = 1.8e - 15), doxorubicin (*P* = 1.45e - 18), paclitaxel (*P* = 3.08e - 28), and sunitinib (*P* = 3.73e - 05) (**Fig. 5E**), demonstrating its robust drug-predictive ability. These findings are consistent with the results obtained from TCGA-THCA cohort, which indicated the robust of the multi-omics classification.

CS2 subtype showed multiple immunocyte infiltrations, and the abundance was similar to the results in TCGA-THCA, which supported that the results are reliable (**Fig. 6A**). We mainly focus on the activated CD8 T cell and Th1 for their unique role in anti-PD-1 or anti-PD-L1 therapy. Responding patients generally showed increased CD8 T cell and Th1 cell infiltrations after treatment with anti-PD-1 as compared with non-responders^45^. We reproduced the SubMap analysis to select the responders to immunotherapy, and the results confirmed CS2 subtype was the potential responder to anti-PD-1 or anti-PD-L1 therapy (**Fig. 6B-C**).

**Figure 6.**
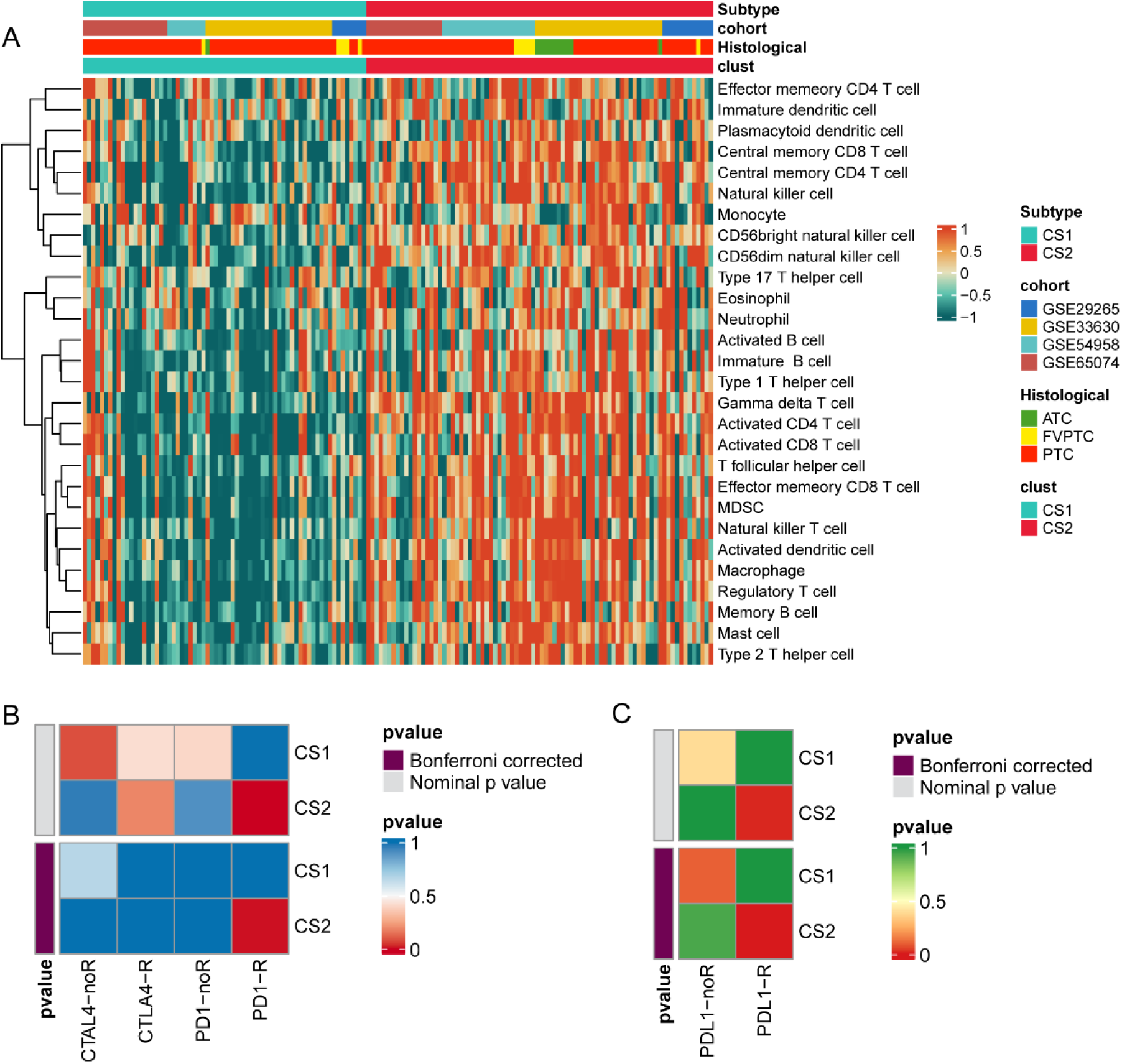
CS1 and CS2 subtypes showed distinct immunocyte infiltration landscapes and different responses to immunotherapy. (A) Distinct immunocyte infiltration landscape in GEO-combined cohort. (B) SubMap analysis showed the responders to anti-PD-1 or anti-CTLA-4 therapy, the dark red square represented the positive group. (C) SubMap analysis revealed the responders to anti-PD-L1 therapy.

### Dimensionality reduction for THCA molecular typing system

Building upon earlier investigations, we have discerned two distinct molecular subtypes of thyroid carcinoma. To streamline the clinical application of this classification system, a dimensionality reduction was undertaken. In layman’s terms, pivotal genes were pinpointed using Variable Selection via Random Forests to achieve molecular typing with a minimal genetic array. We commenced by selecting genes exhibiting a log2 fold change exceeding 2.5 between the two subtypes, applying the “varSelRF” package in R for the reduction process. Subsequently, five genes—CXCL17, LCN2, MUC1, SERPINA1, and SLC34A2—were identified. These genes manifested significantly higher expression in the CS2 subtype compared to CS1 (all P < 0.05, **Figure 7A**). Utilizing these five genes facilitated the distinction between CS1 and CS2 across all participating cohorts with remarkable success: TCGA-THCA (Kappa = 1, P < 0.001), GSE22965 (Kappa = 0.8, P < 0.001), GSE33630 (Kappa = 0.6, P < 0.001), GSE54958 (Kappa = 0.675, P < 0.001), and GSE65074 (Kappa = 0.780, P < 0.001) (**Figure 7B**). The predictive model based on these genes demonstrated excellent efficacy in identifying THCA molecular subtypes (TCGA-THCA, AUC: 1; GSE22965, AUC: 0.979; GSE33630, AUC: 0.776; GSE54958, AUC: 0.975; GSE65074, AUC: 0.970; **Figure 7C**).

**Figure 7:**
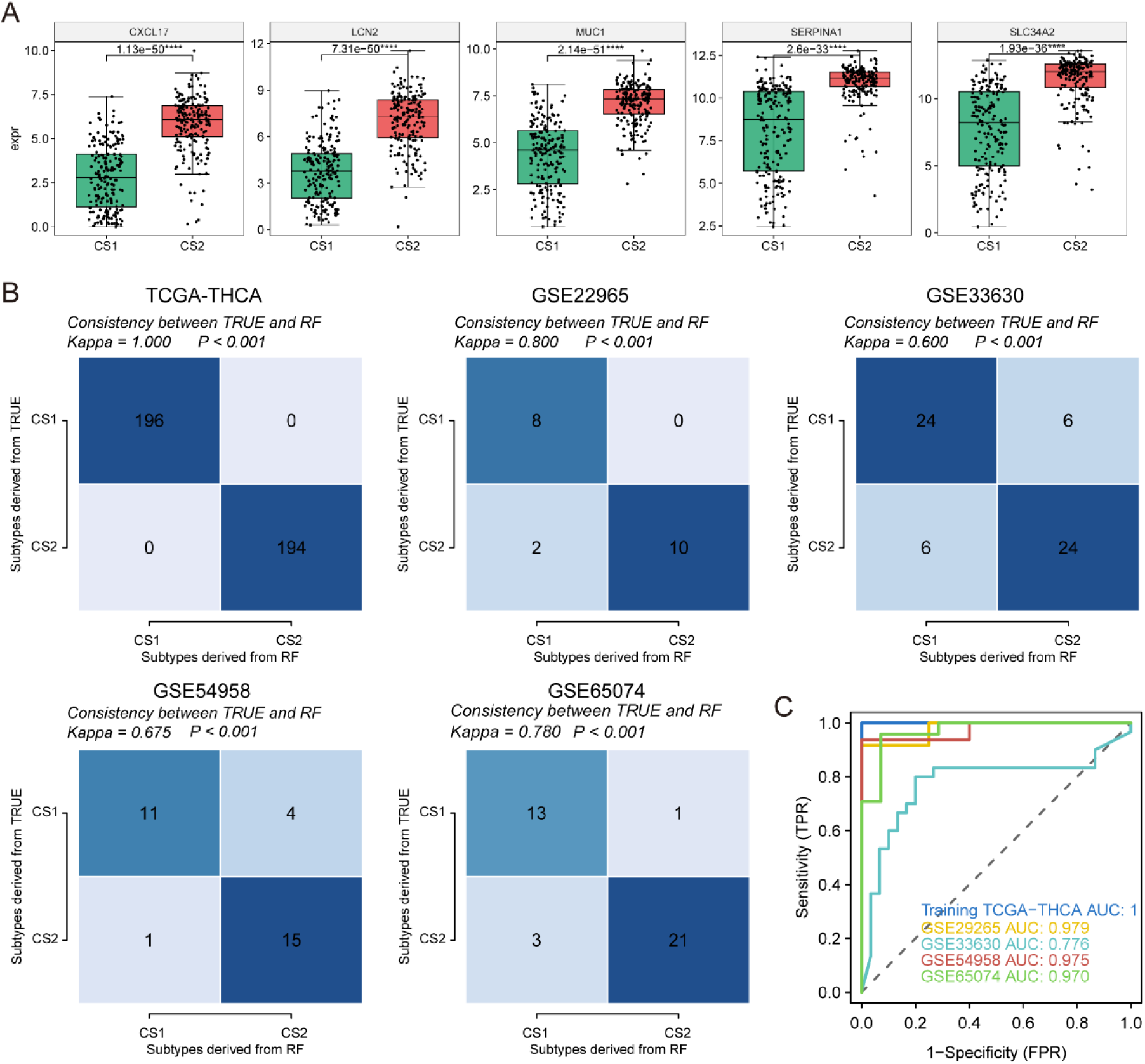
Dimensionality reduction and molecular typing validation for thyroid carcinoma. (A) Expression comparison of the five genes identified for molecular typing. (B) Validation of the molecular typing efficiency across multiple cohorts. (C) Receiver operating characteristic (ROC) curves showcasing the predictive model’s efficacy in different cohorts.

Further validation of CXCL17’s discriminatory role involved analyzing 24 paired tumor and adjacent normal tissues from surgically treated thyroid carcinoma patients through immunohistochemical staining (**Figure 8A**). It was observed that CXCL17 expression was significantly elevated in tumor tissues compared to normal counterparts (Paired T-test, P = 0.013, **Figure 8B**). Moreover, samples with tumor diameters of 1cm or greater exhibited a substantial increase in CXCL17 expression relative to those with smaller diameters (T-test, P = 0.0089, **Figure 8C**). These findings suggest that heightened CXCL17 expression is closely linked to tumorigenesis and advanced disease stages, corroborating our previous analysis which associated elevated CXCL17 levels with the poorer-prognosis CS2 subgroup.

**Figure 8:**
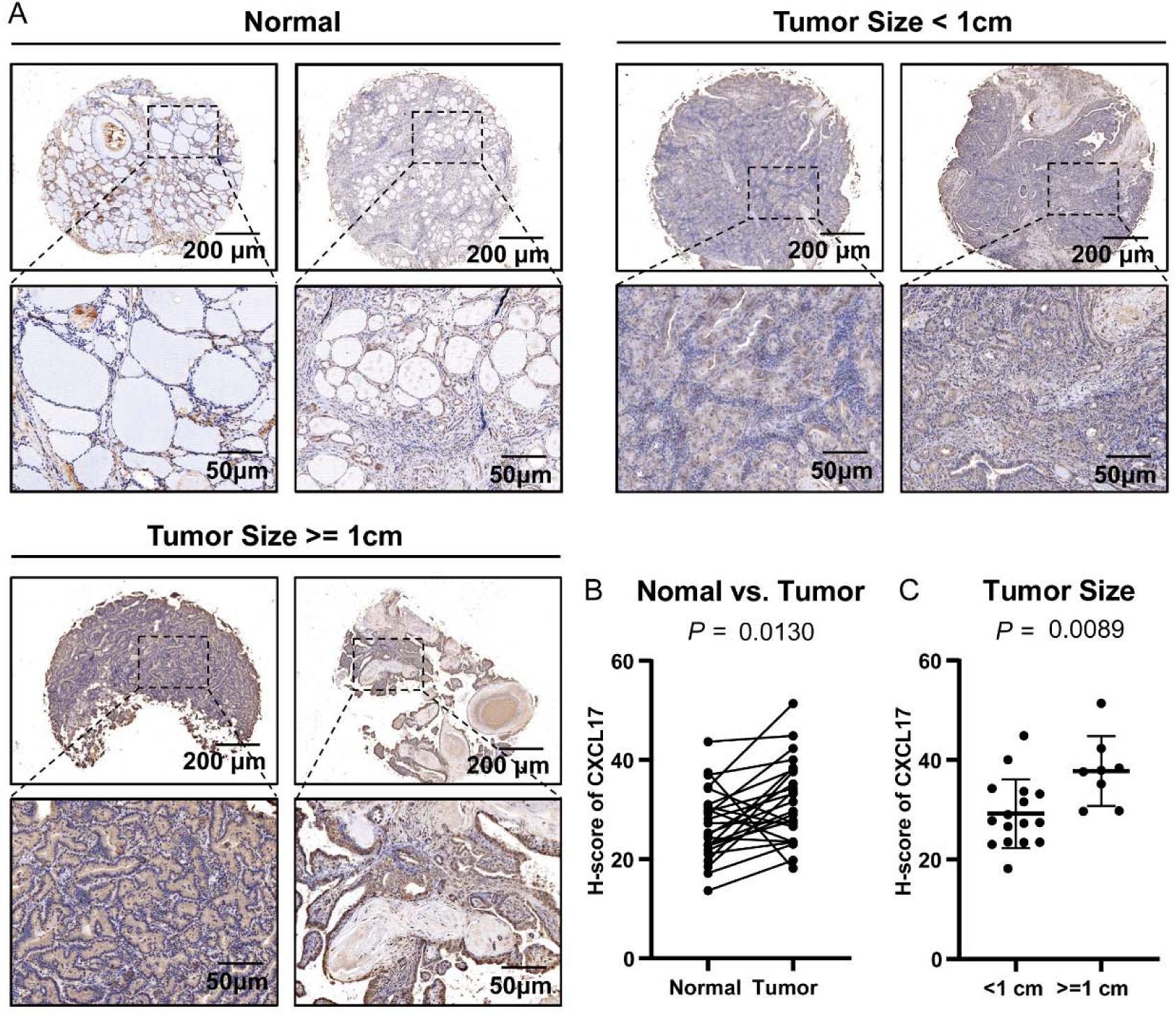
Clinical validation of CXCL17 expression in thyroid carcinoma. (A) Immunohistochemical staining of CXCL17 in paired tumor and adjacent normal thyroid tissue samples. (B) Quantification of CXCL17 expression levels among paired tumor and adjacent normal tissues. (C) Quantification of CXCL17 expression levels among tumor tissue ≥1cm and < 1cm subgroups.

## Discussion

Our comprehensive multi-omics analysis of thyroid cancer (TC) has led to significant insights into the molecular subtypes and their clinical relevance. The incidence and prevalence of TC, particularly among women in China, underscore the importance of this research. The WHO’s 5th edition classification, based on molecular characteristics and aggressiveness, aligns with our findings of distinct molecular subtypes within TC.

The application of the TCGA-THCA dataset and additional cohorts from the GEO database solidifies the identification of two prognostic-relevant subtypes, CS1 and CS2, which show marked differences in progression-free survival (PFS) and clinical outcomes. Our findings highlight the importance of genetic alterations, such as TMB and CNA, in understanding the biology of these subtypes. Notably, CS2 is characterized by a higher tumor mutation burden (TMB) and cytolytic activity (CYT) score, indicative of an aggressive phenotype with a potentially higher immunogenic profile.

The evaluation of immune infiltration further delineates the two subtypes, with CS2 showing a higher infiltration of immune cells and upregulation of PD-1/PD-L1, suggesting a potential responsiveness to immunotherapies targeting these checkpoints. This is particularly relevant given the current landscape of cancer therapy, where immunotherapy plays a pivotal role. The link between the multi-omics subtypes and the established BRAF-RAS classification not only validates our subtyping but also enhances the understanding of TC’s molecular underpinnings.

The activation of distinct pathways, such as cell proliferation and immune-related pathways in CS2, and oxidative phosphorylation in CS1, provides a potential explanation for the differences in prognosis and chemotherapy response. Importantly, these pathway activations offer a molecular basis for the observed drug sensitivities, suggesting personalized therapeutic strategies could be more effective. For instance, CS2’s sensitivity to drugs like cisplatin and doxorubicin aligns with its high proliferation rate and immune activation, while CS1’s sensitivity to bicalutamide and FH535 reflects its distinct metabolic profile.

The validation across multiple cohorts underscores the robustness of our findings and the potential for these molecular subtypes to guide clinical decision-making. Moreover, the reduction to a core set of genes for molecular typing promises a more streamlined, cost-effective approach to patient stratification in clinical settings. The strong correlation between CXCL17 expression and tumor presence, particularly in larger tumors, suggests it may serve as a viable biomarker for the poorer-prognosis CS2 subtype.

## Conclusion

This study offers a detailed characterization of TC subtypes through a multi-omics lens, providing valuable insights into their biological behavior, clinical outcomes, and response to therapy. Our research reinforces the potential of precision medicine in thyroid cancer, advocating for a more targeted approach to treatment based on molecular subtyping. Future research should focus on the clinical validation of these subtypes and their utility in guiding therapeutic decisions to improve patient outcomes.

## Data Availability

All data used in this work are available upon request from the corresponding author.

---

**Figure S1.** Combined raw expression data of GSE29265, GSE33630, GSE54958 and GSE65074 cohorts, and the combined data after removing batch effect.

**Figure S2.** Speculation regarding the appropriate clustering number.

**Figure S3.** The top 100 specific genes with the highest expression in CS1 and CS2 subtypes.

